# Impact of nonpharmaceutical governmental strategies for prevention and control of COVID-19 in São Paulo State, Brazil

**DOI:** 10.1101/2020.08.23.20180273

**Authors:** Cristiane Ravagnani Fortaleza, Thomas Nogueira Vilches, Gabriel Berg de Almeida, Claudia Pio Ferreira, Rejane Maria Tommasini Grotto, Raul Borges Guimarães, Lenice do Rosário de Souza, Carlos Magno Castelo Branco Fortaleza

## Abstract

Interrupted time series analyses (ITSA) were performed to measure the impact of social distancing policies (instituted 22/03/2020) and subsequent mandatory masking in the community (instituted 04/05/2020) on the incidence and effective reproductive number (Rt) of COVID-19 in São Paulo State, Brazil. Overall, the impact of social distancing both on incidence and Rt was greater than the incremental effect of mandatory masking. Those findings may reflect either a small impact of face masking or the loosening of social distancing after mandatory use of masks.

---

São Paulo state reported the first case of COVID-19 in Brazil, on 25 February 2020 [1]. While this manuscript is being written (as of 23 August 2020), the number of laboratory-confirmed cases in Brazil approaches 3 million, with circa 115 thousand deaths (https://covid.saude.gov.br/). Circa 20.5% of both cases and deaths occurred in São Paulo, Brazil’s most populous state (44 million inhabitants). Geographic modeling[2] and ecological analyses[3] documented patterns of disease spread from the metropolitan area, where 39 cities harbor 21 million inhabitants, to the remaining 606 municipalities located in the inner areas of the state.

To contain the spread of SARS-Cov-2, The government of São Paulo decreed the closure of non-essential services and the restriction of public transport from 22 March. Even though evidence for the effectiveness of that measure in preventing deaths and intensive care units (ICU) admissions was reported[4], data on its impact in the inner areas of the state is still lacking. Furthermore, a second decree from May 4^th^ made the use of face masks in community settings mandatory.

Out study was a natural experiment using ecological approach. We aimed at analyzing the impact of both control measures cited above on daily incidence rates (per 100,000 inhabitants) and daily estimates of the effective reproductive number (R_t_) of laboratory-confirmed COVID-19 cases in the metropolitan and inner areas of São Paulo State. Daily incidence from 1 March through 4 July were obtained from São Paulo State Epidemiological Surveillance Center database (http://www.cve.saude.sp.gov.br), and population data for year 2020 was recovered from the São Paulo State Foundation for Data Analysis (SEADE, https://www.seade.gov.br/). The time series of cases was used to estimate R_t_, as described by Wallinga & Lipsitch[5].

Interrupted time series (ITS) analysis^6^ was performed using STATA 14 (Statacorp, College Station, TX), with models that included both successive interventions (social distancing [#1] and universal masking [#2]). Those models fitted linear trends for each period (pre-intervention #1, between interventions, and post-intervention #2). Both immediate impact and sustained changes in trends were assessed.

Results are presented in **Table 1** and **Figure 1**. Social distancing had immediate beneficial impact on incidence in all the state, but trend changes were significant only for inner municipalities. As symmetric result was detected for R_t_: overall immediate impact, but sustained trend changed only in the metropolitan area. The incremental impact of universal masking was not immediate, but a significant change downwards in incidence trends was generally detected. As for the R_t_, only an immediate impact in inner municipalities was statistically significant.

**Table 1.**
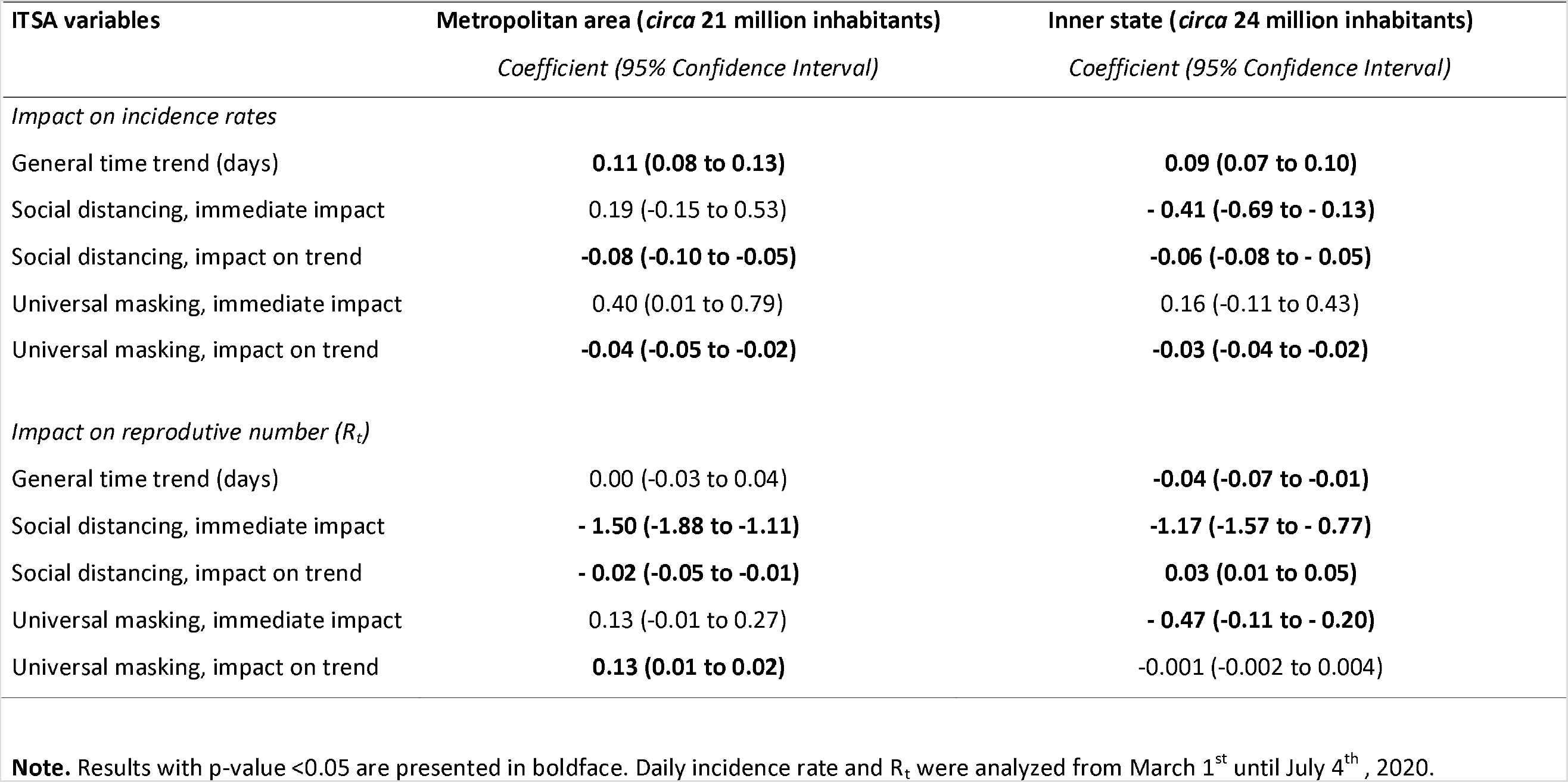
Interrupted time series analysis (ITSA) of the impact of governmental interventions for social distancing (March 22^n^, 2020) and mandatory universal masking (May 4^th^, 2020) on trends of incidence and effective reproduction number (R_t_) of laboratory-confirmed COVID-19 cases in the metropolitan area and inner municipalities of Sao Paulo State, Brazil.

**Figure 1.**
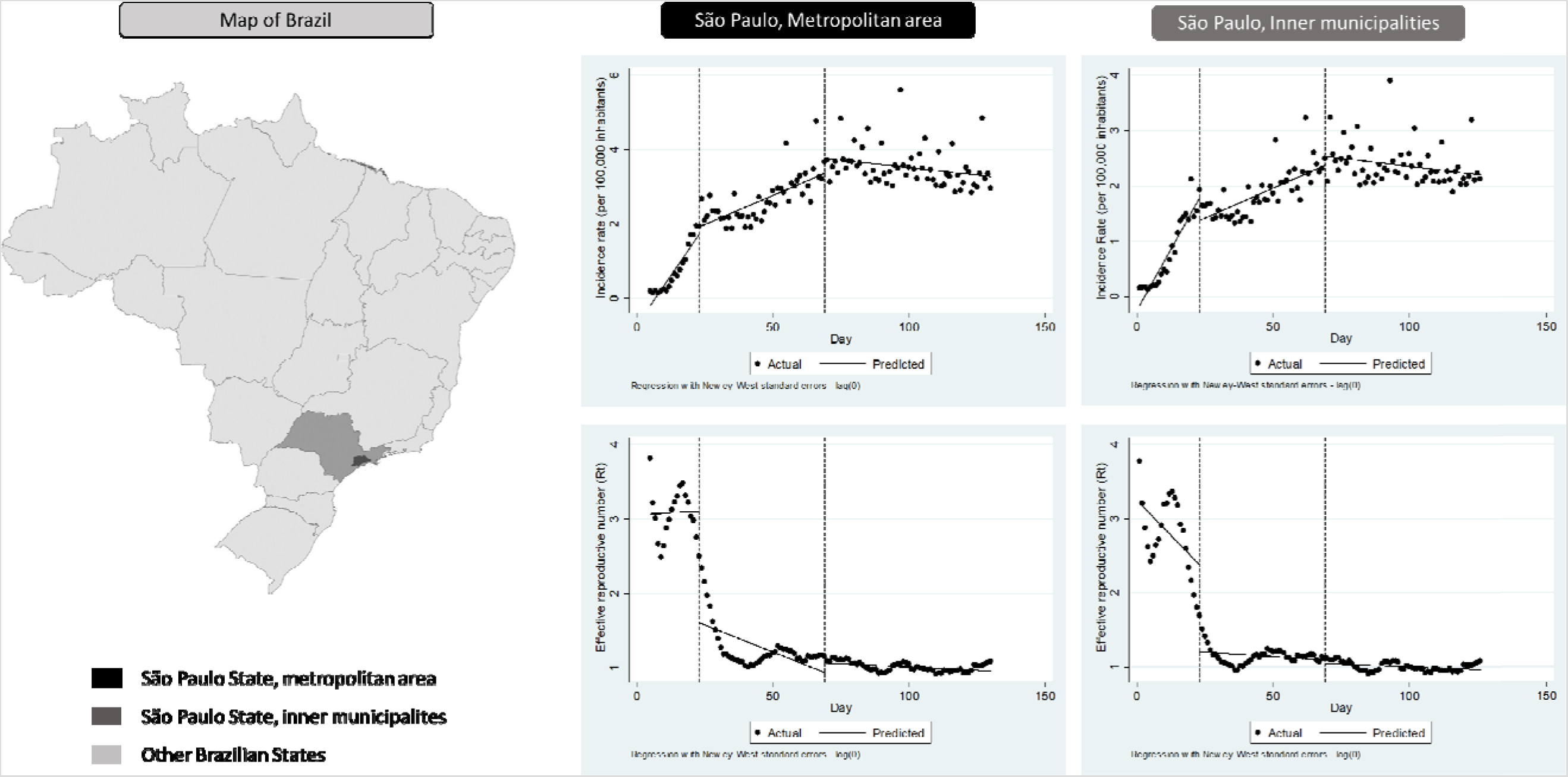
Graphical presentation of the impact of governmental interventions for social distancing and mandatory universal masking on trends of incidence and effective reproduction number (R_t_) of laboratory-confirmed COVID-19 cases in the metropolitan area and inner municipalities of São Paulo State, Brazil. **Note**. The two dashed vertical lines in the graphs represent, respectively, government decrees of social restriction (22 March) and un iversal use of masks in the community (4 May).

São Paulo state did not experience a lockdown in the strict sense of the word. Instead, the March 22^nd^ decree attempted to induce social distancing by closing non-essential services (e.g., stores) and restricting public transport (subway, metropolitan railways, and buses). ITS analysis demonstrates a variable but generally beneficial impact of that measure. This is in agreement with previous reports and modeling predictions[4,7]. The incremental benefit of mandatory universal masking was subtle, with impact on incidence rates but not on daily R_t_ series. This finding contrasts with recent reports from the US [8].

As with all analyzes of natural experiments, our study is subject to confounding bias. The most important is the gradual loosening of restrictive policies (the so-called São Paulo Plan, available at https://www.saopaulo.sp.gov.br/planosp/). Briefly, those measures, initiated in 1 June 2020 were slowly and heterogeneously applied. The State was divided in 22 administrative areas, which could open 0 to 40% of non-essential services, depending on standards of COVID-19 incidence and mortality, availability and occupation of Intensive Care Unit beds.

Municipalities were also allowed to implement additional restrictive measures, as aspect which increased the heterogeneity of the preventive policies. Recommendations changed every two weeks, depending on novel epidemiological data. To assess that slowly applied, geographically heterogeneous and time-varying strategy was beyond the scope of this study. Also, the continuous rise in population mobility (as detected from movement of mobile phones and reported in the São Paulo Government homepage, https://www.saopaulo.sp.gov.br/coronavirus/) was not included in our models, since we had access only to incomplete data. Also, the increasing availability of laboratory testing may have influenced our findings. However, as previously reported, that increase was greater during the first months of pandemics in Brazil, so they were expected to underestimate the impact of the first intervention [9]. Even though those aspects might confound the impact of nonpharmaceutical measures, they are real life phenomena inherent to studies of effectiveness (as opposed to efficacy).

Another limitation of our study was its ecological approach. Obviously, we did not assess the impact of staying at home or using cloth masks on individual risks of being infected by SARS-Cov-2. Our main objective was to study the collective impact of governmental policies.

A systematic review of nonpharmaceutical measures to prevent COVID-19, based solely on observational studies, concluded that social distancing, use of masks and eye protection were beneficial [10]. Unfortunately, those results are limited by the heterogeneity of settings (healthcare and community) and of interventions. The analysis of benefits from use of masks is particularly challenging. Previous studies point to an expected greater efficacy of N95 respirators and surgical masks than that of cloth and/or homemade masks. It is worth noting that a recent systematic review with meta-analysis or randomized clinical trials found general protective impact of face masks [11]. However, protection was greater in healthcare settings than in the community. Also, the included studies assessed influenza and other respiratory viruses, and most of them tested surgical masks. Therefore, inferences have been largely based on analogy, and the benefits of cloth masks are far from straightforward.

In conclusion, we found that governmental strategies based on nonpharmaceutical intervention were generally effective in slowing the evolution of pandemics in São Paulo State, Brazil. The effectiveness was greater for the first intervention (social distancing), with some incremental impact of mandatory use of face masks. Those findings may reflect either a small impact of face masking or the loosening of social distancing after mandatory use of masks. Either way, they contribute for directing public policies against COVID-19 in Brazil and other countries, in a period when the world is still far from achieving control of the current pandemics.

## Data Availability

Data on which the research was based is uploaded in two supplementary files

## Acknowledgements

All authors state that they have no conflict of interest regarding this study. The authors received no funding.

## Data availability statement

All data used for our analysis is available in two supplementary files uploaded with the manuscript.

